# Prophylactic antibiotics to prevent ventilator associated pneumonia in adults with acute brain injury who are invasively ventilated in the ICU: A protocol for a systematic review and meta-analysis

**DOI:** 10.1101/2023.06.05.23290875

**Authors:** Laura Hailstone, Kate Hadley-Brown, Roisin Devane, Josh Davis, Naomi Hammond, Qiang Li, Ed Litton, John Myburgh, Joseph Santos, Ian Seppelt, Steven Y.C. Tong, Andrew Udy, Balasubramanian Venkatesh, Paul Young, Anthony Delaney

## Abstract

**Background:** Acute brain injury due to conditions such as trauma, subarachnoid haemorrhage, stroke or hypoxic-ischaemic encephalopathy, is a major public health issue. Lower respiratory tract infections and ventilator associated pneumonia (VAP), are common in patients who require invasive mechanical ventilation after suffering an acute brain injury, and may have potentially deleterious consequences such as fever, hypoxaemia, and hypotension, excessive pulmonary secretions and sputum plugging. These physiological disturbances may contribute to secondary brain injury and potentially to adverse clinical outcomes. Parenteral antibiotics given soon after the commencement of invasive mechanical ventilation may prevent the development of VAP and potentially reduce the associated adverse consequences, although there is conflicting evidence from randomised clinical trials (RCTs).

Therefore, we plan to conduct a systematic review and meta-analysis to test the hypothesis that, in adults with acute brain injury who are invasively ventilated in the Intensive Care Unit (ICU), administration of prophylactic parenteral antibiotics, compared with a matched placebo or usual care, reduces the occurrence of mortality as well as ventilator associated pneumonia and other secondary outcomes.

**Methods and analysis:** We will undertake a systematic review and meta-analysis. We will include RCTs that compare the administration of prophylactic antibiotics to placebo or usual care on hospital mortality and other patient-centred outcomes in patients with acute brain injury receiving mechanical ventilation in the ICU. We will perform a search that includes the electronic databases MEDLINE and EMBASE and clinical trial registries. Two reviewers will independently screen titles and abstracts, perform full article reviews and extract study data, with discrepancies resolved by a third reviewer. We will report study characteristics and quantify risk of bias. We will perform random effects meta-analyses to provide pooled estimates that the administration of prophylactic antibiotics is associated with reduced hospital mortality and a reduced incidence of ventilator associated pneumonia, as well as other outcomes. We will evaluate overall certainty of evidence using the Grading of Recommendations Assessment, Development, and Evaluation framework.

**Conclusion:** This systematic review and prospective meta-analysis will provide clinicians with an overview of current evidence regarding the association between the administration of prophylactic antibiotics in patients with acute brain injury receiving invasive mechanical ventilation and mortality, the incidence of ventilator associated pneumonia and other clinical outcomes.

**PROSPERO registration:** CRD 42023424732

## Introduction

Acute brain injury due to conditions such as trauma, subarachnoid haemorrhage, stroke or hypoxic-ischaemic encephalopathy, is a major public health issue.(1) While a significant proportion of the mortality and long term-morbidity following acute brain injury is determined by the nature and severity of the primary insult, as well as patient factors such as age and co-morbidity, a major focus of critical care management is to prevent secondary injury brain insults such as hypoxia, hypotension, and raised intracranial pressure,(2, 3) as well as preventing secondary complications that commonly follow acute brain injury.(4, 5)

Lower respiratory tract infections and ventilator associated pneumonia (VAP), referred to hereafter as VAP, are common in patients who require invasive mechanical ventilation after suffering an acute brain injury. Patients with acute brain injury are at particular risk of developing VAP due to the reduced level of consciousness and impairment of upper airway reflexes related to the severity of the primary brain injury.(6) The development of VAP in patients with severe acute brain injury may have potentially deleterious consequences. Fever, hypoxaemia, and hypotension, resulting from sepsis related to pulmonary infection can contribute to secondary brain injury.(2) Excessive pulmonary secretions can lead to sputum plugging and coughing, which may result in elevations of arterial carbon dioxide causing elevated intracranial pressure.(2) Pulmonary infection can lead to delayed extubation and failed extubation, which are known to be associated with adverse outcomes.(7, 8)

Parenteral antibiotics may prevent the development of VAP and potentially reduce the associated adverse consequences. A prior systematic review has addressed this issue.(9) This study included only 2 randomised clinical trials, and found a reduction in the incidence of VAP, and a reduction in Intensive Care Unit (ICU) length of stay, but no effect on mortality. Longer term neurological or functional outcomes were not reported. Subsequent randomised clinical trials in populations with acute severe brain injury have been reported without a clear signal of benefit or harm.(10, 11, 12)

Therefore, we plan to conduct a systematic review and meta-analysis to test the hypothesis that, in adults with acute brain injury who are invasively ventilated in the ICU, administration of prophylactic parenteral antibiotics, compared with a matched placebo or usual care, reduces the occurrence of mortality as well as ventilator associated pneumonia and other secondary outcomes.

## Methods

We will conduct a systematic review of randomised clinical trials with meta-analysis in accordance with the recommendations of the Cochrane Handbook of Systematic Reviews of Interventions(13) and will report the findings as per the Preferred Reporting Items for Systematic Review and Meta-Analysis (PRISMA) statement.(14) This systematic review has been submitted for registration on the International Prospective Register of Systematic Reviews (CRD42023424732)

### Eligibility Criteria

#### Study types

We will include randomised clinical trials. There will be no restriction on publication status, language, nor year of publication.

#### Population

We will include trials in which the population is adults with acute brain injuries due to traumatic brain injury, ischaemic stroke, intracerebral haemorrhage, subarachnoid haemorrhage, status epilepticus, and suspected hypoxic ischaemic encephalopathy following resuscitation from cardiac arrest who are undergoing invasive mechanical ventilation in an ICU.

#### Intervention

We will include trials in which the intervention is the administration of prophylactic parenteral antibiotics within 48 hours of initiation of invasive mechanical ventilation. There will be no restriction on the type of antibiotic or on duration of administration of the intervention.

#### Comparison

We will include trials in which the comparison group is either allocated to placebo or standard care.

#### Outcomes

We will include studies that report any of the outcomes specified for this review.

### Exclusion criteria

- Trials that solely use oral antiseptic agents
- Trials where antibiotics are administered as part of a strategy of selective decontamination of the digestive tract.

### Information sources

We will perform a search of the electronic databases MEDLINE using the PubMed interface as well as EMBASE, and the Cochrane Central Register of Clinical Trials using the OVID interface, to identify reports of clinical trials and conference abstracts. We will search clinical trial registries through the World Health Organisation international clinical trials registry platform and clinicaltrials.gov. We will search for unpublished trials by searching the PubMed indexed pre-print servers, as well as by contacting experts in the field. We will manually search reference lists of included studies and other reviews and perform forward and backward citation screening for all included trials.

### Search strategy

We will develop our search strategy in alignment with the Peer Review of Electronic Search Strategies (PRESS) guideline statement.(15) We will use a combination of search terms to identify studies that include populations with traumatic brain injury, ischaemic stroke, intracerebral haemorrhage, subarachnoid haemorrhage, status epilepticus, and suspected hypoxic ischaemic encephalopathy following resuscitation from cardiac arrest, mechanical ventilation and antibiotic prophylaxis and combine them with sensitive filters to identify randomised clinical trials.(16, 17) The full electronic search strategy is shown in the Supplement.

### Data management and selection process

Study records identified by the search will be downloaded into a reference management system software.(18) Duplicate records will be removed. Two authors will independently screen the titles and abstracts of identified study records to identify potentially eligible studies. We will retrieve full text manuscripts of studies that are deemed potentially eligible by either reviewer for detailed review.

### Selection process

Two authors will independently apply the eligibility criteria to all potentially eligible studies. Differences will be resolved by discussion or by a third reviewer. Reasons for exclusion will be recorded and presented in a PRISMA flow diagram.(14)

### Data collection process

Data extraction forms will be developed, and pilot tested. Data will be extracted from the included studies onto specific data collection forms. Data extraction will be performed in duplicate, with discrepancies resolved by discussion or by a third reviewer. Where data are not available in the primary manuscript, we will attempt to obtain additional data by contacting the authors of included studies, or by obtaining data from previously published sources.

### Data items

We will extract data regarding study characteristics (including first author, year of publication, number of participants, number of sites), details of the included ICUs (e.g., medical, surgical, trauma, mixed general ICU) and participants (including age distribution, sex distribution, nature of head injury, duration of ventilation at baseline, severity of illness), details of the interventions (antibiotic agents, dose and duration) as well as details of the comparison group (placebo or usual care), use of topical antiseptics, use of VAP bundles, definition of VAP. Outcome data by group will be collected including VAP incidence, hospital mortality, ICU and hospital length of stay, duration of ventilation during the study, reintubation, and neurological function at 6-month outcomes.

### Risk-of-Bias assessment

Two review authors, with no affiliation with any of the included RCTs, will independently assess the risk of bias for each included study. We will use all publicly available reports of trials, including published trial protocols and statistical analysis plans to assess risk of bias. Disagreements will be resolved by discussion, involving a third independent assessor if needed. We will use the Cochrane Risk of Bias tool.(19) Details regarding the risk of bias assessments will be reported in the electronic supplement. We will present a ‘risk of bias summary’ figure, depicting each risk of bias item for each included study.

### Outcomes

#### Primary

The primary outcome will be hospital mortality (or the closest timepoint to hospital mortality available)

#### Secondary

The Secondary outcomes will be:

- The incidence of ventilator associated pneumonia (as defined in the included trials)
- ICU length of stay
- Duration of mechanical ventilation
- Reintubation
- Hospital length of stay
- Neurological function at 6 months
- Detection of antimicrobial resistant organisms

### Data analyses

The main analyses will be performed on all included trials regardless of risk of bias. For binary outcomes, we will report risk ratios (RR) while for continuous outcomes mean differences (MD) or standardised MD. We will perform random effects meta-analyses using the Sidik-Jonkman method(20) to provide pooled estimates, along with the 95% confidence intervals.(21) As a sensitivity analysis, we will perform fixed effects meta-analysis.(22) We will try to obtain missing outcome data from the original study authors, we will not impute any missing data. Analyses will be conducted on an intention to treat basis. All pooled results will be presented in the form of forest plots. We will also undertake a trial sequential analysis using standard methods.(23)

### Assessment of heterogeneity

We will assess quantitative heterogeneity by reporting the proportion of total variability due to heterogeneity rather than due to sampling error (I^2^).(24)

### Assessment for small study/publication bias

We will evaluate small-study effects by visual assessment of the contour-enhanced funnel plots(25) and formal Egger’s regression test.

### Predefined subgroup analysis

We will conduct three subgroup analyses of the primary outcome.

- We hypothesise that the effect of prophylactic antibiotics will be less in those with hypoxic ischaemic encephalopathy compared to other causes of acute severe brain injury.
- We hypothesise that the effect of prophylactic antibiotics will be more pronounced in trials in which the prophylactic antibiotics were administered for more than 48 hours compared to trials in which the prophylactic antibiotics were administered for 48 hours or less.
- We hypothesis that the effect of prophylactic antibiotics will be more pronounced in trials in which the prophylactic antibiotics were commenced within 12 hours of initiation of mechanical ventilation compared to trials with longer durations in which the prophylactic antibiotics were to be commenced.

The credibility of any subgroup analysis will be assessed using the Instrument to assess the Credibility of Effect Modification Analyses (ICEMAN) in randomized clinical trials and meta-analyses.(26)

## Confidence in cumulative evidence

We will use the Grading of Recommendations Assessment, Development, and Evaluation (GRADE)(27) approach to assess the overall certainty of evidence that the administration of prophylactic antibiotics compared with placebo or usual care improves outcome for each primary and secondary outcome measure and present the results in a ‘summary of findings’ table(28) including plain language summary.(29) The certainty of evidence and our confidence in the effect-estimates and strength of treatment recommendation will be evaluated by the discussion with all authors and be based on the methods in the Cochrane Handbook of Systematic Reviews and updated GRADE working group recommendations.(29, 30) The overall certainty of evidence will be rated “high”, “moderate”, “low” or “very low” for each outcome

## Ethics and dissemination

This review does not require ethical approval as this is a systematic review of published studies. We plan to present the results of the systematic review at national and international scientific meetings and will prepare a manuscript for submission to a peer reviewed journal.

## Discussion and limitations

This systematic review and meta-analysis will provide the most up-to-date synthesis of evidence regarding the effect of the administration of prophylactic parenteral antibiotics on the incidence of ventilator associated pneumonia in invasively ventilated adults with acute brain injuries in the ICU. We acknowledge that there will be limitations to the proposed systematic review, including that eligible studies are anticipated to be heterogeneous in nature due to variations in the interventions given, trial design, and variation in treatment of the comparison groups. In addition, the strength of a systematic review and meta-analysis relies in part on the strength of included studies, and therefore may be limited due to the paucity of high quality randomised clinical trials in this area.

## Data Availability

All data produced in the present study are available upon reasonable request to the authors

## Supplement

### Search strategy

#### CENTRAL

EBM Reviews - Cochrane Central Register of Controlled Trials <March 2023>

1. antibiotic prophylaxis.mp. or exp antibiotic prophylaxis/
2. prophylactic antibiotics.mp. or exp antibiotic prophylaxis/
3. infection control.mp. or exp infection control/
4. antibiotics.mp. or exp antibiotic agent/
5. healthcare associated pneumonia.mp. or exp health care associated pneumonia/
6. ventilator associated pneumonia.mp. or exp ventilator associated pneumonia/ or exp artificial ventilation/
7. mechanical ventilation.mp. or exp artificial ventilation/
8. artificial respiration.mp. or exp artificial ventilation/
9. subarachnoid haemorrhage.mp. or exp subarachnoid hemorrhage/
10. status epilepticus.mp. or exp status epilepticus/
11. craniocerebral trauma.mp. or exp head injury/
12. traumatic brain injury.mp. or exp traumatic brain injury/
13. stroke.mp. or exp cerebrovascular accident/
14. ischaemic stroke.mp. or exp ischemic stroke/
15. intracranial haemorrhage.mp. or exp brain hemorrhage/
16. Post cardiac arrest syndrome.mp. or exp post-cardiac arrest syndrome/
17. cardiac arrest.mp. or exp Heart Arrest/
18. hypoxic ischaemic encephalopathy.mp. or exp hypoxic ischemic encephalopathy/
19. brain hypoxia.mp. or exp brain hypoxia/
20. subdural haematoma.mp. or exp subdural hematoma/
21. subdural haemorrhage.mp. or exp subdural hematoma/
22. exp coma/ or coma.mp.
23. 1 or 2 or 3 or 4
24. 5 or 6 or 7 or 8
25. 9 or 10 or 11 or 12 or 13 or 14 or 15 or 16 or 17 or 18 or 19 or 20 or 21 or 22
26. 23 and 24 and 25

#### EMBASE

Database: Embase <1974 to 2023 April 19>

Search Strategy:

_________________________________________

1. antibiotic prophylaxis.mp. or exp antibiotic prophylaxis/
2. prophylactic antibiotics.mp. or exp antibiotic prophylaxis/
3. infection control.mp. or exp infection control/
4. antibiotics.mp. or exp antibiotic agent
5. healthcare associated pneumonia.mp. or exp health care associated pneumonia/
6. ventilator associated pneumonia.mp. or exp ventilator associated pneumonia/ or exp artificial ventilation
7. mechanical ventilation.mp. or exp artificial ventilation/
8. artificial respiration.mp. or exp artificial ventilation/
9. subarachnoid haemorrhage.mp. or exp subarachnoid hemorrhage/
10. status epilepticus.mp. or exp status epilepticus/
11. craniocerebral trauma.mp. or exp head injury/
12. traumatic brain injury.mp. or exp traumatic brain injury/
13. stroke.mp. or exp cerebrovascular accident/
14. ischaemic stroke.mp. or exp ischemic stroke/
15. intracranial haemorrhage.mp. or exp brain hemorrhage/
16. Post cardiac arrest syndrome.mp. or exp post-cardiac arrest syndrome/
17. cardiac arrest.mp. or exp Heart Arrest/
18. hypoxic ischaemic encephalopathy.mp. or exp hypoxic ischemic encephalopathy/
19. brain hypoxia.mp. or exp brain hypoxia/
20. subdural haematoma.mp. or exp subdural hematoma/
21. subdural haemorrhage.mp. or exp subdural hematoma/
22. exp coma/ or coma.mp.
23. 1 or 2 or 3 or 4
24. 5 or 6 or 7 or 8
25. 9 or 10 or 11 or 12 or 13 or 14 or 15 or 16 or 17 or 18 or 19 or 20 or 21 or 22
26. random:.tw.
27. clinical trial:.mp.
28. exp health care quality/
29. 26 or 27 or 28 6616078
30. 23 and 24 and 25 and 29

#### Pubmed

((((((antibiotic prophylaxis) OR (prophylactic antibiotics)) OR (infection control)) OR (antibiotics)) AND ((((healthcare associated pneumonia) OR (ventilator associated pneumonia)) OR (mechanical ventilation)) OR (artificial respiration))) AND ((((((((((((((coma) OR (subdural haematoma)) OR (subdural haemorrhage)) OR (brain hypoxia)) OR (hypoxic ischaemic encephalopathy)) OR (coma post cardiac arrest)) OR (post cardiac arrest syndrome)) OR (intracerebral haemorrhage)) OR (ischaemic stroke)) OR (stroke)) OR (traumatic brain injury)) OR (craniocerebral trauma)) OR (subarachnoid haemorrhage)) OR (status epilepticus))) AND ((“clinical”[Title/Abstract] AND “trial”[Title/Abstract]) OR “clinical trials as topic”[MeSH Terms] OR “clinical trial”[Publication Type] OR “random*”[Title/Abstract] OR “random allocation”[MeSH Terms] OR “therapeutic use”[MeSH Subheading])

